# Digital maturity and its determinants in General Practice: a cross-sectional study in 20 countries

**DOI:** 10.1101/2022.08.23.22278753

**Authors:** Fábia Teixeira, Edmond Li, Liliana Laranjo, Claire Collins, Greg Irving, Maria Jose Fernandez, Josip Car, Mehmet Ungan, Davorina Petek, Robert Hoffman, Azeem Majeed, Katarzyna Nessler, Heidrun Lingner, Geronimo Jimenez, Ara Darzi, Cristina Jácome, Ana Luísa Neves

**Author notes:** **Correspondence:** Ana Luísa Neves.

## Abstract

**Background:** The extension to which digital technologies are employed to promote the delivery of high-quality healthcare is known as Digital Maturity. Individuals’ and systems’ digital maturity are both necessary to ensure a successful, scalable and sustainable digital transformation in healthcare. Digital maturity in primary care has been scarcely evaluated.

**Objectives:** This study assessed the digital maturity - as a whole and in its dimensions - in General Practice and evaluated how participants’ demographic characteristics, practice characteristics and features of Electronic Health Records (EHRs) use are associated with digital maturity.

**Methods:** General Practitioners (GPs) across 20 countries completed an online questionnaire between June and September 2020. Demographic data, practice characteristics and features of EHRs use were collected. Digital maturity was evaluated through a framework built upon usage, resources and ability (divided in this study in its collective and individual components), interoperability, general evaluation methods and impact of digital technologies. Each dimension was rated as 1 or 0. The digital maturity score is the sum of the six dimensions and ranges from 0 to 6 (maximum digital maturity). Multivariable linear regression was used to model the total score, while multivariable logistic regression was used to model the probability of meeting each dimension of the score.

**Results:** 1,600 GPs (61% female, 68% Europeans) participated. GPs had a median digital maturity of 4 (P25-P75: 3-5). Positive associations with digital maturity were found for: being male (B=0.18 [95%CI 0.01;0.36]), use of EHRs for longer periods (B=0.45 [95%CI 0.35;0.54]) and higher frequencies of access to EHRs (B=0.33 [95%CI 0.17;0.48]). Practising in a rural setting was negatively associated with digital maturity (B=-0.25 [95%CI −0.43;-0.08]). Usage (90%) was the most acknowledged dimension while interoperability (47%) and use of best practice general evaluation methods (28%) were the least. Shorter durations of EHRs use were negatively associated with all digital maturity dimensions (aOR from 0.09 to 0.77).

**Conclusions:** Our study demonstrated notable factors that impact digital maturity and exposed discrepancies in digital transformation across healthcare settings. It provides a roadmap for policymakers to develop more efficacious interventions to hasten and take the best advantage of digital transformation in General Practice.

## Introduction

Digital technologies have revolutionised many aspects of modern society and health care is no exception.^1^ Around the world, the onset of the digital transformation has radically changed the primary care landscape ^2,3^ through the widespread computerisation and the digitalisation of personal health information into Electronic Health Records (EHRs). Simultaneously, the dissemination of electronic medical devices,^4^ as well as adoption of systems enabling digital drug prescriptions, referrals, billing, scheduling tests and appointments, are also major contributors to this change.^5^ Advances in digital technologies can also be seen from the proliferation of implantable devices which offer real time monitoring of physiological parameters, to telemedicine^1^ and mobile health - the use of mobile devices to improve health outcomes.^6^ This already ongoing digital transition has been further accelerated by the COVID-19 pandemic.^1,7^

From facilitating communication between providers to improving prevention, achieving early diagnosis and providing timely treatments, digital technologies have a tremendous potential to improve health care delivery,^8,9^ however they have not yet played a major role among efforts to improve primary healthcare.^10^ Nonetheless, the relevance of digital technologies keeps growing in primary care as governments approaches to this sector continue to move towards the use of more collaborative systems.^3^ The extension to which digital technologies are employed to promote the delivery of high quality healthcare is known as Digital Maturity and it is an emerging concept across developed health care systems.^11^ The digital maturity of health professionals and systems is necessary to ensure a successful, scalable and sustainable digital transformation. ^12,14^

More than modernisation of medical resources, digital transformation is a complex multidimensional process^6^ and therefore digital maturity, as any care intervention, needs to be rigorously evaluated and monitored to ensure successful implementation.^11^ While there have been studies focused on the assessment of digital maturity in secondary care,^15,16^ research is also needed in primary care practice.^3^ To our best knowledge, digital maturity in primary care has not been previously evaluated.

This study assesses the digital maturity - as a whole and in its dimensions - in general practice across 20 countries and evaluates if the characteristics of participants or clinical practices, as well as features of EHR adoption, are associated with digital maturity. Our hypothesis is that the characteristics described above can affect digital maturity. The identification of such factors may contribute to developing more efficacious digital transformation implementation strategies worldwide.

## Methods

### Study Design and Setting

This is a cross-sectional study, which used an online questionnaire completed by GPs. It was granted ethical approval from the Imperial College Research Ethics Committee (Reference 20IC5956), which oversees health-related research with human participants. The study adheres to the STrengthening the Reporting of OBservational studies in Epidemiology (STROBE) guideline for cross-sectional studies. The research was conducted by a primary care consortium (inSIGHT Research Group) which gathers health professionals from 20 countries (Australia, Brazil, Canada, Chile, Colombia, Croatia, Finland, France, Germany, Ireland, Israel, Italy, Poland, Portugal, Slovenia, Spain, Sweden, Turkey, the United Kingdom, and the United States).

### Participants

Participants were eligible if they were GPs working in the countries above between March and September 2020.

### Data Collection

Participants were recruited between June and September 2020. Investigators at the Patient Safety Translational Research Centre and Department of Primary Care and Public Health at Imperial College London constructed the questionnaire. It was piloted by the national leads of the 20 inSIGHT Research Group associate countries in May 2020 and edited for national, cultural or organisational adaptations. The questionnaire was originally developed in English, and was translated to French, German, Italian, Portuguese and Spanish by national leads to stimulate higher participation. It was provided to participants through Qualtrics. National leads invited GPs working in their country to take part in the questionnaire via email and through social media channels, such as Facebook and LinkedIn.^17^

Demographic data (gender, age and country), practice features (setting, number of hours of clinical work per week, number of years of experience as GP and involvement in teaching activities) and characteristics of access to EHRs (availability of EHRs, duration and frequency of use) were collected. Digital maturity was assessed using the digital maturity framework developed by Flott et al which is built upon usage, resources and ability, interoperability, general evaluation method, and impact.^11^ The following statements were used to assess the items described:

- Usage: “*Most healthcare providers in our practice use the digital system*”
- Resources & ability (organisational): “*Our organisation is ready to use the digital system correctly*”
- Resources & ability (individual): “*We have the individual abilities needed to use the digital system correctly*”
- Interoperability: “*Our digital system has the capability to communicate across services or with other systems*”
- General evaluation methodology: “*We have best practice digital maturity evaluation methods in place*”
- Impact: “Our system has a positive impact in terms of outcomes for patients, structure, process or finance”.

Each of the six statements corresponds to one dimension. All dimensions were evaluated by the participant as one of the following options: agree, neutral or disagree. For each participant, a score of agreement with the above dimensions addressing digital maturity was calculated. Whenever the participant expressed agreement with one dimension, 1 point was granted. The score is the sum of the 6 dimensions and ranges from 0 to 6 (maximum digital maturity).

### Data Analysis

The sample size is superior to the total number of responses calculated to be needed to provide a confidence level of 95% and a margin of error of 5% (901), according to the published protocol reported elsewhere.17

All participants, even those in which some parameters were missing, were used in the analysis. Countries were categorised as European (Finland, France, Germany, Ireland, Italy, Poland, Portugal, Slovenia, Spain, Sweden and the United Kingdom) and Non-European (remaining). The variable “Setting of practice” was split into “Rural” and “Urban”. The option “Prefer not to answer” in the questions regarding age, gender and involvement in teaching activities were treated as missing information.

The normal distribution of each continuous variable was assessed by Kolmogorov-Smirnov tests. Quantitative data were analysed using absolute and relative frequencies for categorical variables and continuous variables with skewed distribution using median and interquartile range. Univariate linear regression was performed to determine the characteristics associated with the digital maturity score (i.e., gender, age, country, years of experience as GP, hours of clinical work per week, involvement in teaching activities, rural setting of practice, urban setting of practice, access to EHRs, duration and frequency of use of EHRs). All independent variables associated with digital maturity score with a P<.12 were included in the first multivariable model iteration. The variables for multivariable analysis were chosen through the stepwise method. Unstandardized coefficients (B) and 95% Confidence Intervals were calculated. The models were evaluated using p values and coefficients of determination (R^2^). Similarly, univariate binomial logistic regressions were used to identify characteristics possibly predicting the outcome (0=neutral/disagree, 1=agree) of each of the 6 components of the digital maturity score usage, collective resources and ability, individual resources and ability, interoperability, general evaluation methods and impact. Characteristics with P<.12 at univariate analysis were used in a multivariable logistic regression. The final model was obtained using a forward conditional regression. Adjusted Odds Ratio and 95% Confidence Intervals (aOR (95% CI)) were calculated. The models were evaluated using Hosmer Lemeshow tests and Nagelkerke’s R-square. Data were analysed using IBM SPSS Statistics 26.0 (IBM Corporation, Armonk, NY, USA).

## Results

### Participants characteristics

A total of 1600 GPs were enrolled, mostly female (61%; n=976) aged between 30 to 39 years old (33%; n=530) and practising in European countries (68%; n=1,081). Most of them had more than 20 years of experience as a GP (31%; n=431), worked a median of 36 hours per week (P25-P75: 28-40), in an urban setting (73%, n=1,354) and were involved in teaching activities (64%; n=1,017). Most of them had access to EHRs (95%, n=1,523) and were using it every day (91%, n=1,379) for more than 10 years (55%, n=838). The characteristics of the participants are summarised in Table 1.

**Table 1.**
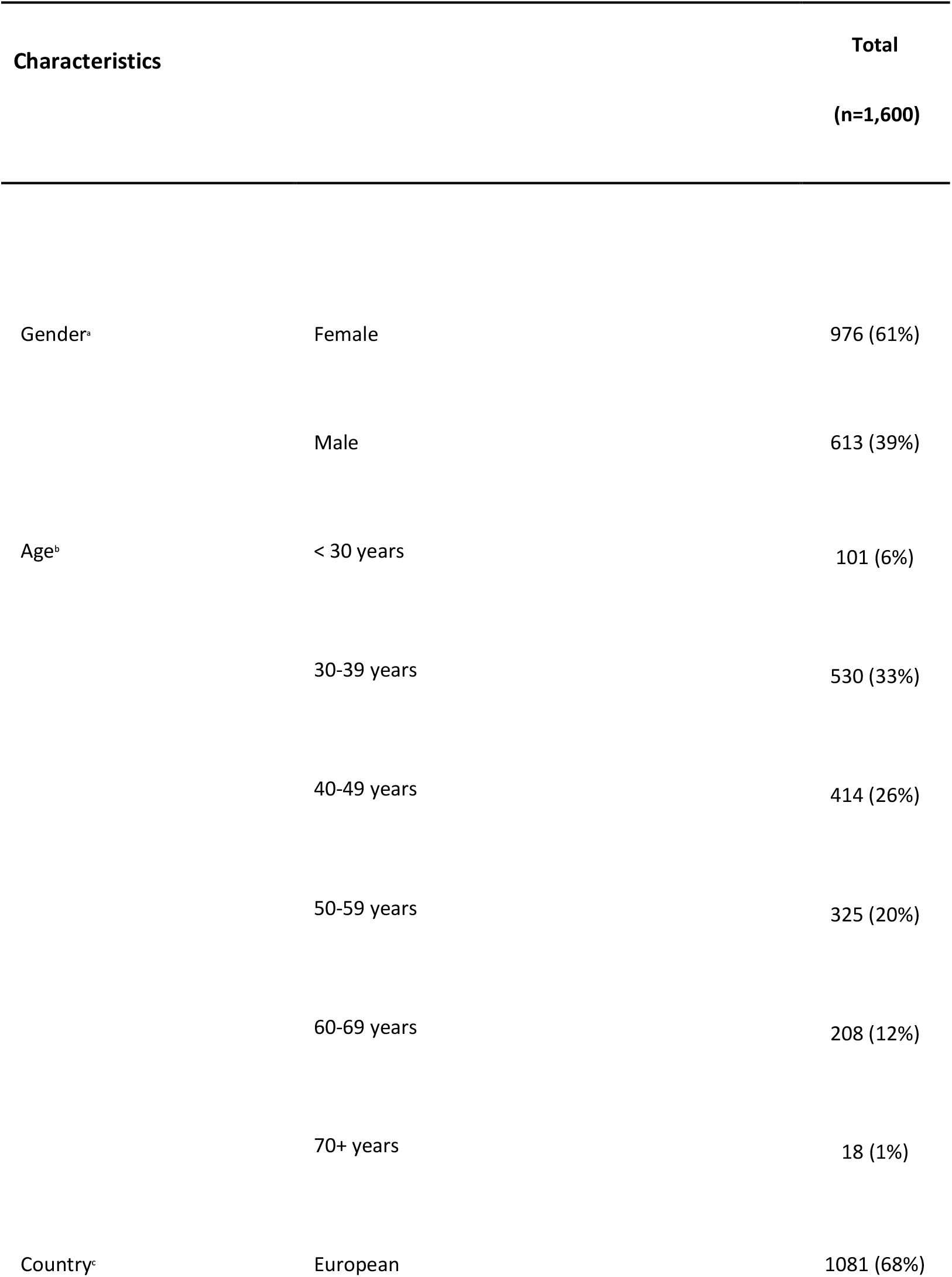
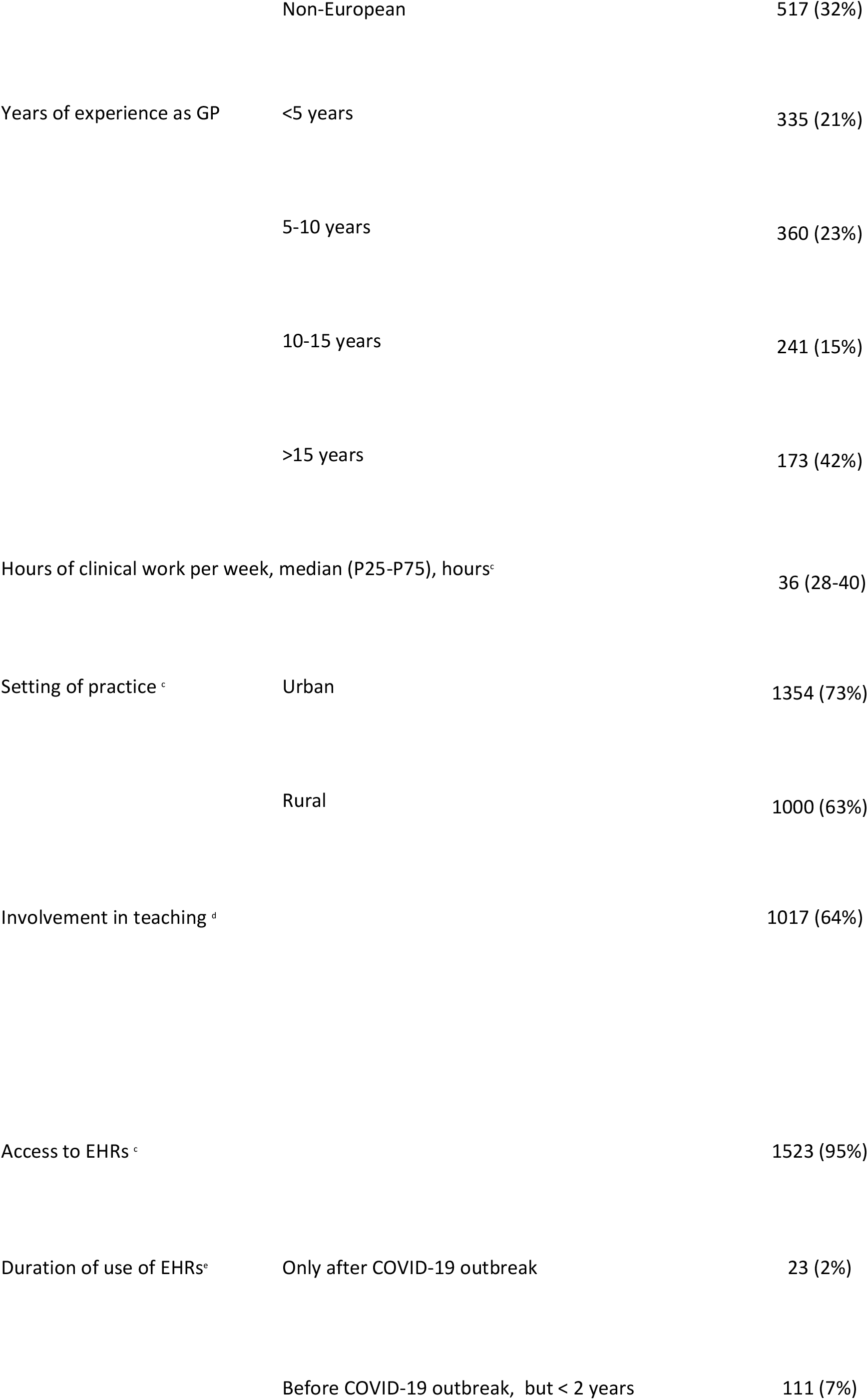
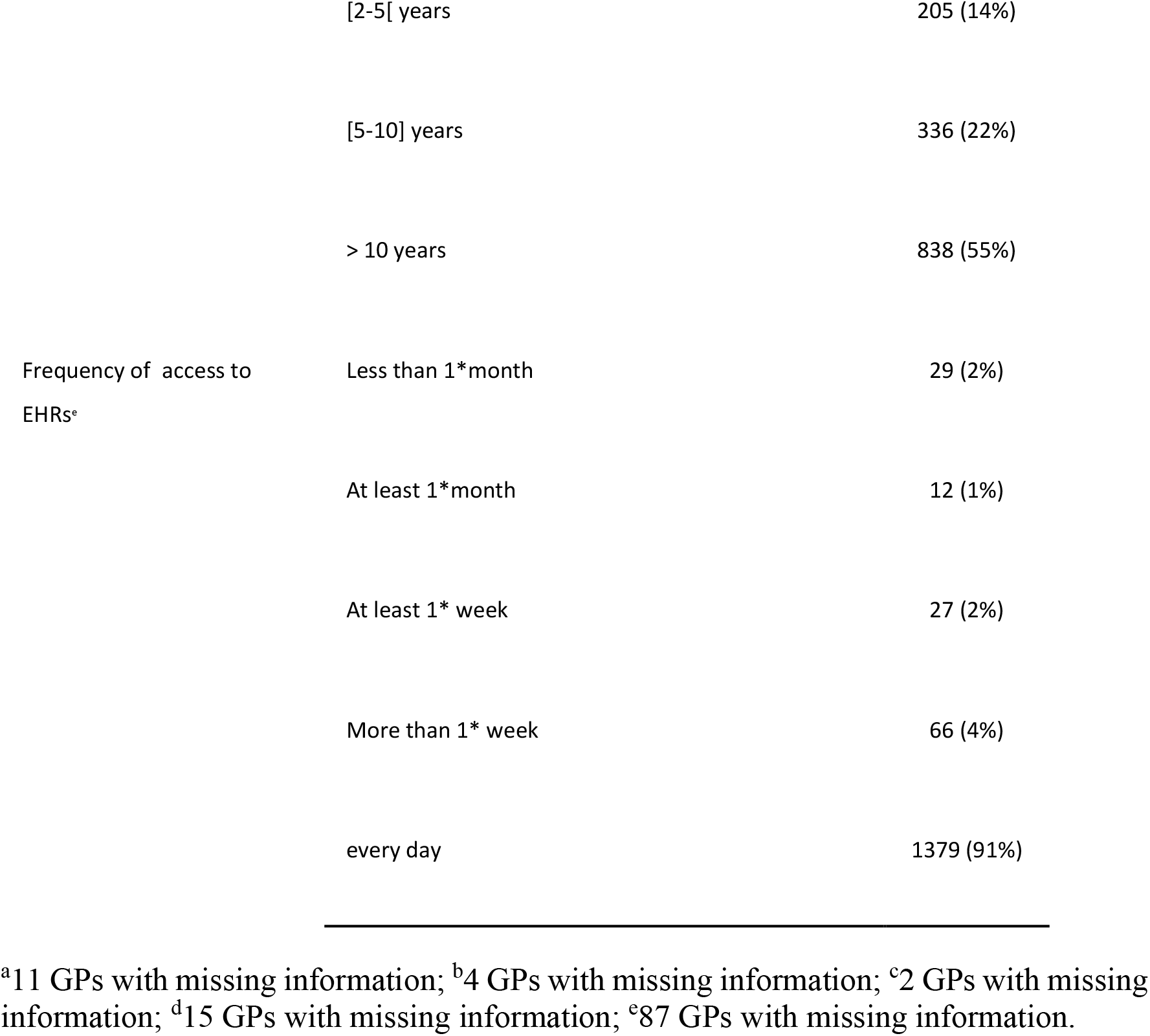
Participants characteristics (n=1,600). Unless otherwise indicated, values are displayed in n (%). GP-General Practitioner, P25-P75 - Percentile 25 to Percentile 75, EHRs - Electronic Health Records

### Digital maturity and participants’ characteristics

Participants had a median digital maturity score of 4 (3-5). The highest 3 levels of the score accounted for almost 60% of the answers. Among the six dimensions, usage registered the highest percentage of agreement (90%, n=1,209), followed by collective and individual resources and ability (80%, n=1073 and 77%, n=1035, respectively), impact (59%, n=788) and interoperability (47%, n=633). Best practice general evaluation methods registered the lowest scores of agreement (28%, n=380). A significant multivariable linear regression model explained the digital maturity score (R2 = 11%, *P*<0.001). Being male was associated with a higher digital maturity score (B=0.18 [95%CI 0.01;0.36]), while practising in a rural setting was inversely associated with it (B=-0.25 [95%CI −0.43;-0.08]). Additionally, longer duration and higher frequency of use of EHRs were also associated with a higher digital maturity score (B=0.45 [95%CI 0.35;0.54], B=0.33 [95%CI 0.17;0.48], respectively). A detailed overview of the model is provided in Table 2.

**Table 2.** Univariate and Multivariable linear regression models to explain the digital maturity score. Ref - Reference, B- unstandardized regression coefficient, 95% CI - 95% confidence interval, GP-General Practitioner, EHRs - Electronic Health Record

| Characteristics | Univariate analysis |  | Multivariable analysis |  |
| --- | --- | --- | --- | --- |
|  | B [95%CI] | P value | B [95%CI] | P value |
| Gender (Ref=Female) | 0.27 [0.08;0.45] | 0.005 | 0.18 [0.01;0.36] | 0.042 |
| Age | 0.18 [0.11; 0.26] | <0.001 |  |  |
| Country (Ref= Non-European) | 0.26 [0.07;0.45] | 0.008 |  |  |
| Years of experience as GP | 0.21 [0.13;0.28] | <0.001 |  |  |
| Hours of clinical work per week | -0.01 [-0.01;0.01] | 0.864 |  |  |
| Rural setting of practice | -0.15 [-0.34;0.40] | 0.114 | -0.25 [-0.43;-0.08] | 0.005 |
| Urban setting of practice | -0.03 [-0.28;0.22] | 0.797 |  |  |
| Involvement in teaching activities | 0.19 [-0.01;0.38] | 0.056 |  |  |
| Access to EHRs | 0.28 [-0.18;0.74] | 0.229 |  |  |
| Duration of use of EHRs | 0.53 [0.44;0.61] | <0.001 | 0.45 [0.35; 0.54] | <0.001 |
| Frequency of access to EHRs | 0.57 [0.42;0.72] | <0.001 | 0.33 [0.17;0.48] | <0.001 |

### Individual dimensions of digital maturity and participants’ characteristics

Unadjusted ORs estimating the association between the characteristics of the participants and each of the 6 dimensions of the digital maturity are presented in Table 3. Urban setting of practice was not associated with any dimension, while duration of use of EHRs was associated with all of them.

Adjusted ORs (aORs) represent the multivariable analysis of the predictors of each dimension and are summarised in Table 4. The models explained 19% of the variance of usage, 13% of collective resources and ability, 6% of individual resources and ability, 7% of interoperability, 4% of general evaluation methods and 6% of impact. Hosmer Lemeshow tests showed that the models adequately fitted the data (*P*=.713, *P*= .983, *P*=.276, *P*=.554, *P*=.981, *P*=.956, respectively).

#### Usage

GPs were less likely to use digital systems if they were using EHRs for a shorter period of time (aOR from 0.09 to 0.52) when compared to GPs accessing them for more than 10 years. Lower frequencies of access to EHRs were also associated with lower odds of use of the digital systems (aOR from 0.18 to 0.43) when compared to accessing them every day. On the other hand, GPs who had been working as it for less years had higher odds of using the digital systems (aOR from 1.58 to2.42) when compared to being a GP for more than 15 years. The number of hours GPs worked in a week were negatively associated with usage of digital technologies (aOR= 0.99 [0.98;1.00]).

#### Collective Resources and ability

When compared to GPs accessing EHRs for more than 10 years, GPs who started accessing them later were less likely to express having collective resources and abilities (aOR from 0.14 to 0.54), as well as GPs who access EHRs less frequently (aOR from 0.39 to 0.85) when compared to GPs accessing them every day.

#### Individual Resources and ability

Being male was positively associated with reporting individual resources and ability (aOR 1.33 [95%CI 1.00;1.80]), while practising in a rural setting was negatively associated with it (aOR 0.67 [95%CI 0.51;0.88]). GPs who started accessing EHRs more recently were less likely to acknowledge individual resources and abilities (aOR from 0.47 to 0.77), when compared to GPs accessing them for more than 10 years. GPs who accessed EHRs less frequently were also less likely to acknowledge individual resources and ability (aOR from 0.20 to 0.55) when compared to GPs accessing them every day.

#### Interoperability

In comparison with non-European GPs, Europeans were more likely to identify interoperability in the digital system they used (aOR= 1.42 [1.11;1.80]). In contrast, GPs who started accessing EHRs more recently were less likely to identify interoperability (aOR from 0.28 to 0.51) than those who have been accessing them for more than 10 years.

#### General Evaluation Methods

Being European was associated with lower odds of practising the best digital systems evaluation methods (aOR 0.68 [0.52;0.88]). Likewise, having started to access EHRs more recently was associated with lower odds of having best practice evaluation methods in place (aOR from 0.27 to 0.65).

#### Impact

Males had higher odds of reporting digital system’s impact (aOR1.35), as well as younger GPs (aOR 3.41 to 5.30) when compared to being 70 or more years old. On the other hand, in comparison with GPs who started to access EHRs over than 10 years ago, GPs who started accessing them more recently were associated with lower odds of recognizing impact of the digital systems they used (aOR from 0.33to 0.62). Similarly, when compared to GPs with every day access to EHRs, GPs with less frequent accesses were less likely to identify impact as an asset of the digital systems (aOR from 0.16 to 0.86).

## Discussion

### Principal Findings

GPs had an overall good digital maturity score. While the overall usage was the most acknowledged dimension of the digital maturity evaluation framework (90%), interoperability (47%) and use of best practice evaluation methods (28%) were the dimensions receiving a lower score, highlighting the potential for improvement in these areas.

Being male, longer duration of use of EHRs and higher frequency of access to EHRs were positively associated with self-reported digital maturity, foreseeing their function as predictors. Practising at a rural setting was negatively associated with digital maturity, exposing discrepancies on the digital transformation across settings. No significant associations were found with age, country, years of experience as GP, hours of clinical work per week, urban setting of practice, involvement in teaching activities, and access to EHRs.

All six dimensions of digital maturity might be explained by distinct characteristics, with shorter durations of use of EHRs being negatively associated with all of them.

### Comparison with Previous Literature

There has been an increase in the number of studies focused on developing digital maturity evaluation tools.^18,19^ Although a considerable amount of research on this topic has been recently issued, to our knowledge, there are no studies reporting the usage of such tools in primary care.

The World Health Organization has already recognised investment in resources, strategies for maximising impact, standardised evaluation metrics and interoperability of systems as key to the success of digital transformation.^20^ Interestingly, we found interoperability and general evaluation models to be the most prevalent shortcomings of digital systems maturity. Previous evidence regarding the determinants of digital health transformation in integrated care in Europe reported that although interoperability relevance is greatly understood, the maturity of its implementation is currently rather poor,^21^ which is consistent with our findings. However, comparisons between studies should be cautious as different tools were used to assess digital maturity.

Previous studies on the analysis of digital maturity determinants in secondary care focused on understanding whether availability of resources was related to digital maturity. In hospitals, investment in hardware and software was positively associated with higher levels of digital maturity.^22^ The effect of demographic factors, practice characteristics and adoption of EHRs features on digital maturity is lacking in literature.

Zaresani A and Scott A have suggested that physicians who used digital health technology were more likely to be male.^22^ In the present study, being male was positively associated with digital maturity, but this information should be carefully considered due to the possibility of the existence of other factors playing a role in this association. For example, we can hypothesise that this relation might be explained by the chance men are more prone to self-report digital maturity than women.

Gheorghiu B and Hagens S conducted a study in Canada to study the adoption of interoperable EHRs across different jurisdictions. They concluded that jurisdictions where physicians accessed interoperable EHRs more often were also the ones where its use had been happening for longer periods of time. They used the frequency of end users’ access to EHRs as a method of gauging the systems’ maturity^23^ Indeed, in the present study, GPs accessing EHRs more frequently were associated not only with higher overall digital maturity, but also with better scores on usage, collective and individual resources and abilities and impact. The duration of use of EHRs was also associated with better overall digital maturity and with each of its six dimensions.

Regarding the clinical practice in rural areas, this was negatively associated with the maturity of digital systems. Although there was a lack of evidence specifically exploring the impact of the practice setting in the digital maturity of health systems, it is reported in the literature that rural areas remain left behind in terms of broadband and other digital connectivity, not to mention in terms of digital adoption and skills.^24^

### Strengths and Limitations

This study has several strengths. To the best of our knowledge, it is the first study focusing on the evaluation of digital maturity in primary care and the exploitation of its determinants. Participants were GPs working from 20 different countries worldwide, with diversified resource management policies in primary care. A comprehensive set of participants’ demographic characteristics, practice characteristics and features of EHRs adoption was collected and analysed, which allowed us to explore their role in digital maturity.

However, this study has some limitations that should be acknowledged. It is based on an non validated questionnaire, which gives no guarantees that the collected variables are truly measuring digital maturity. The questionnaire was disseminated online via email and social media channels and therefore a potential selection bias cannot be excluded. For example, we can hypothesise that GPs that were more prone to answer the online questionnaire were those working with higher digital maturity. This can possibly explain that 55% of the participants were using EHRs for more than 10 years and 91% were accessing them every day. Additionally, the lack of translation of this questionnaire to the official languages of all 20 inSIGHT Research Group member countries might have presented an obstacle to its enrollment in certain countries. Nevertheless, this data collection methodology enabled us to gather data from 20 countries in a short period of time, proving it to be prompt, economical, and safe to use. Due to its cross-sectional design, this study only enabled us to assess digital maturity during a specific period. It would be important to reproduce this online questionnaire in the future, to allow deductions on the digital maturity temporal evolution to be made.

Additionally, the framework developed by Flott K *et al* was used to evaluate digital maturity at the primary care level only. This choice was made since the focus of our work was in fact general practice. Future studies should consider the utilisation of the entire framework in its 4 levels (home, community, primary and secondary care) since the evaluation of the digital maturity of health services is dependent on a sector wide patient understanding.^11^

Finally, most GPs included in this study were female (61%), European (68%), involved in teaching activities (64%). Therefore, attempts to generalise these findings to populations with different characteristics need to be cautious.

### Conclusions

This is the first international study performed in general practice providing important results for putting into practice in different levels. This work generates evidence on the level of digital maturity in primary care. It exposes interoperability and best practice evaluation methods as the most prevalent digital maturity shortcomings in primary care, which represents greater potential for improvement in these two dimensions. Our results disclose a negative association between practising general medicine in a rural setting and the level of digital maturity, highlighting discrepancies across various healthcare settings which can slow overall digital transformation.

Therefore, our findings provide a roadmap for stakeholders in digital health, mainly to policymakers, to develop increasingly effective strategies to hasten and take the best advantage of the ongoing digital transformation in General Practice.

## Data Availability

The subsets of the database analyzed for this study are available upon reasonable request to the corresponding author.

## Author Contributions

FT, CJ and ALN wrote the first manuscript. All authors reviewed the manuscript and approved the version submitted for publication.

## Acknowledgments

The authors would like to thank all General Practitioners who participated in this study.

