## Supplementary material for "Digital maturity and its determinants in General Practice: a cross-sectional study in 20 countries": Table 4

Table 4 - Multivariable logistic regression models to explain each dimension of the framework: usage, collective resources and ability, individual resources and ability, interoperability, general evaluation and impact. Reference – the category used as reference. OR – Odds Ratio; 95%CI - 95% Confidence Interval; GP- General Practitioner, EHRs - Electronic Health Records

| | **Usage** | | **Collective Resources and ability** | | **Individual Resources and ability** | | **Interoperability** | | **General Evaluation Methods** | | **Impact** | |
|---|---|---|---|---|---|---|---|---|---|---|---|---|
| **Characteristics** | OR [95% CI] | P value | OR [95%CI] | P value | OR [95% CI] | P value | OR [95% CI] | P value | OR [95% CI] | P value | OR [95% CI] | P value |
| **Gender** | | | | | | | | | | | | |
| Male | | | | | 1.33 [1.00;1.80] | 0.047 | | | | | 1.35 [1.07;1.72] | 0.013 |
| Female | Reference | | | | | | | | | | | |
| **Age** | | | | | | | | | | | | |
| < 30 years | | | | | | | | | | | 5.30 [1.58;17.76] | 0.007 |
| 30-39 years | | | | | | | | | | | 3.69 [1.21;11.26] | 0.022 |
| 40-49 years | | | | | | | | | | | 4.58 [1.50;13.97] | 0.007 |

| | | | | | | | | | | | | |
|---|---|---|---|---|---|---|---|---|---|---|---|---|
| 50-59 years | | | | | | | | | | | 3.82 [1.25;11.66] | 0.019 |
| 60-69 years | | | | | | | | | | | 3.41 [1.11;10.48] | 0.032 |
| 70+ years | Reference | | | | | | | | | | | |
| **Country** | | | | | | | | | | | | |
| European | | | | | | | 1.42 [1.11;1.80] | 0.003 | 0.68 [0.52;0.88] | 0.005 | | |
| Non-European | Reference | | | | | | | | | | | |
| **Years of experience as GP** | | | | | | | | | | | | |
| <5 years | 2,42 [1.37;4.38] | 0.003 | | | | | | | | | | |
| 5-10 years | 1.72 [0.97;3.05] | 0.063 | | | | | | | | | | |
| 10-15 years | 1.58 [0.83;3.01] | 0.161 | | | | | | | | | | |
| >15 years | Reference | | | | | | | | | | | |
| **Hours of clinical work per week** | 0.99 [0.98;1.00] | 0.022 | | | | | | | | | | |

| | | | | | | | | | | | | |
|---|---|---|---|---|---|---|---|---|---|---|---|---|
| **Rural setting of practice** | | | | | | | | | | | | |
| Yes | | | | | 0.67 [0.51;0.88] | 0.004 | | | | | | |
| No | Reference | | | | | | | | | | | |
| **Duration of use of EHRs** | | | | | | | | | | | | |
| Only after COVID-19 outbreak | 0.12 [0.04;0.37] | <0.001 | 0.14 [0.05;0.40] | <0.001 | 0.49 [0.17;1.44] | 0.195 | 0.29 [0.10;0.80] | 0.018 | 0.35 [0.10;1.22] | 0.099 | 0.33 [0.11;1.04] | 0.059 |
| Before COVID-19 outbreak. but < 2 years | 0.09 [0.04;0.18] | <0.001 | 0.16 [0.09;0.26] | <0.001 | 0.47 [0.28;0.80] | 0.005 | 0.28 [0.16;0.47] | <0.001 | 0.27 [0.14;0.53] | <0.001 | 0.49 [0.28;0.83] | 0.008 |
| [2-5[ years | 0.17 [0.09; 0.31] | <0.001 | 0.27 [0.19;0.40] | <0.001 | 0.47 [0.33;0.69] | <0.001 | 0.43 [0.30;0.60] | <0.001 | 0.65 [0.31;0.70] | <0.001 | 0.54 [0.37;0.79] | 0.001 |
| [5-10] years | 0.52 [0.29;0.92] | 0.028 | 0.54 [0.37:0.77] | 0.001 | 0.77 [0.55;1.07] | 0.119 | 0.51 [0.39;0.68] | <0.001 | 0.61 [0.45;0.82] | 0.001 | 0.62 [0.46;0.84] | 0.002 |
| > 10 years | Reference | | | | | | | | | | | |
| **Frequency of access to EHRs** | | | | | | | | | | | | |
| Less than 1* month | 0.18 [0.06;0.54] | 0.002 | 0.39 [0.13;1.18] | 0.095 | 0.35 [0.12;1.02] | 0.054 | | | | | 0.16 [0.03;0.75] | 0.020 |

| | | | | | | | | | | | | |
|---|---|---|---|---|---|---|---|---|---|---|---|---|
| At least 1*month | 0.33 [0.08;1.38] | 0.130 | 0.50 [0.13;1.93] | 0.315 | 0.20 [0.05;0.75] | 0.017 | | | | | 0.35 [0.0.9;1.42] | 0.141 |
| At least 1* week | 0.43 [0.16;1.17] | 0.098 | 0.85 [0.32;2.21] | 0.732 | 0.55 [0.21;1.41] | 0.211 | | | | | 0.86 [0.34;2.19] | 0.755 |
| More than 1* week | 0.28 [0.14;0.55] | <0.001 | 0.41 [0.23;0.74] | 0.003 | 0.47 [0.26;0.84] | 0.011 | | | | | 0.66 [0.38;1.16] | 0.153 |
| Everyday | Reference | | | | | | | | | | | |
