## Supplementary material for "Digital maturity and its determinants in General Practice: a cross-sectional study in 20 countries": Table 3

Table 3 - Univariate logistic regression models to explain each dimension of the framework: usage, collective resources and ability, individual resources and ability, interoperability, general evaluation methods and impact. Reference – the category used as reference. OR – Odds Ratio; 95%CI - 95% Confidence Interval; GP- General Practitioner, EHRs - Electronic Health Records

| | **Usage** | | **Collective Resources and Ability** | | **Individual Resources and Ability** | | **Interoperability** | | **General Evaluation Methods** | | **Impact** | |
|---|---|---|---|---|---|---|---|---|---|---|---|---|
| **Characteristics** | OR [95% CI] | P value | OR [95% CI] | P value | OR [95% CI] | P value | OR [95% CI] | P value | OR [95% CI] | P value | OR [95% CI] | P value |
| **Gender** | | | | | | | | | | | | |
| Male | 0.80 [0.55;1.17] | 0.255 | 0.74 [0.55;0.98] | 0.035 | 0.73 [0.56;0.96] | 0.024 | 0.78 [0.63;0.98] | 0.031 | 0.93 [0.73;1.19] | 0.569 | 0.74 [0.59:0.93] | 0.009 |
| Female | Reference | | | | | | | | | | | |
| **Age** | | | | | | | | | | | | |
| < 30 years | 0.71 [0.08;6.27] | 0.761 | 0.16 [0.02;1.27] | 0.083 | 0.19 [0.02;1.54] | 0.120 | 0.65 [0.21;1.99] | 0.454 | 0.56 [0.15;2.03] | 0.376 | 2.67 [0.83;8.55] | 0.099 |
| 30-39 years | 0.55 [0.07;4.25] | 0.564 | 0.21 [0.03;1.61] | 0.133 | 0.19 [0.02;1.42] | 0.105 | 0.69 [0.25;1.95] | 0.486 | 0.82 [0.25;2.62] | 0.732 | 2.31 [0.76;6.86] | 0.133 |
| 40-49 years | -0.81 [0.10;6.36] | 0.838 | 0.44 [0.06;3.43] | 0.433 | 0.30 [0.04;2.29] | 0.243 | 1.08 [0.38;3.04] | 0.887 | 1.36 [0.43;4.38] | 0.603 | 3.50 [1.17;10.48] | 0.025 |
| 50-59 years | 0.64 [0.08;5.05] | 0.672 | 0.32 [0.04;2.48] | 0.275 | 0.25 [0.03;1.94] | 0.185 | 1.47 [0.52;4.17] | 0.468 | 1.23 [0.38;3.96] | 0.734 | 3.17 [1.05;9.52] | 0.040 |

| | | | | | | | | | | | | |
|---|---|---|---|---|---|---|---|---|---|---|---|---|
| 60-69 years | 0.80 [0.10;6.48] | 0.834 | 0.35 [0.05;2.76] | 0.319 | 0.34 [0.04;2.66] | 0.303 | 1.54 [0.53;4.42] | 0.420 | 1.38 [0.42;4.49] | 0.598 | 3.13 [1.03;9.52] | 0.044 |
| 70+ years | Reference | | | | | | | | | | | |
| **Country** | | | | | | | | | | | | |
| European | 1.64 [1.14;2.37] | 0.008 | 1.53 [1.16;2.02] | 0.003 | 1.31 [1.00;1.71] | 0.050 | 1.55 [1.22;1.94] | <0.00 1 | 0.76 [0.59;0.98] | 0.031 | 1.21 [0.96;1.52] | 0.105 |
| Non-European | Reference | | | | | | | | | | | |
| **Years of experience as GP** | | | | | | | | | | | | |
| <5 years | 0.66 [0.41;1.04] | 0.073 | 0.47 [0.33;0.66] | <0.001 | 0.56 [0.40;0.78] | 0.001 | 0.46 [0.34;0.62] | <0.00 1 | 0.57 [0.40;0.80] | 0.001 | 0.75 [0.56;1.01] | 0.060 |
| 5-10 years | 0.94 [0.58;1.53] | 0.940 | 0.88 [0.61;1.27] | 0.491 | 0.85 [0.60;1.19] | 0.336 | 0.53 [0.40;0.71] | <0.00 1 | 0.71 [0.51;0.97] | 0.032 | 0.817 [0.61;1.09] | 0.166 |
| 10-15 years | 1.04 [0.586;1.841] | 0.897 | 0.95 [0.62;1.5] | 0.807 | 0.87 [0.59;1.28] | 0.471 | 0.78 [0.56;1.07] | 0.121 | 1.172 [0.84;1.65] | 0.357 | 0.87 [0.63;1.21] | 0.414 |
| >15 years | Reference | | | | | | | | | | | |
| **Hours of clinical work per week** | 0.99 [0.98;1.00] | 0.065 | 0.99 [0.99;1.00] | 0.099 | 0.999 [0.99;1.01] | 0.580 | 1.00 [0.10;1.01] | 0.505 | 1.00 [0.10;1.01] | 0.307 | 1.00 [1.0;1.01] | 0.459 |
| **Urban Setting of practice** | | | | | | | | | | | | |
| Yes | 0.64 [0.37;1.16] | 0.147 | 1.18 [0.82;1.69] | 0.381 | 1.17 [0.83;1.65] | 0.383 | 0.80 [0.60;1.08] | 0.149 | 1.09 [0.78;1.52] | 0.620 | 0.93 [0.69;1.27] | 0.659 |
| No | Reference | | | | | | | | | | | |

| | | | | | | | | | | | | |
|---|---|---|---|---|---|---|---|---|---|---|---|---|
| **Rural setting of practice** | | | | | | | | | | | | |
| Yes | 1,00 [0.69;1.46] | 0.998 | 0.81 [0.62;1.07] | 0.142 | 0.77 [0.60;1.01] | 0.055 | 1.01 [0.81;1.26] | 0.964 | 0.83 [0.65;1.07] | 0.152 | 0.86 [0.69;1.08] | 0.862 |
| No | Reference | | | | | | | | | | | |
| **Involvement in teaching activities** | | | | | | | | | | | | |
| Yes | 1.07 [0.73;1.58] | 0.718 | 1.23 [0.93;1.64] | 0.144 | 1.08 [0.82;1.42] | 0.574 | 1.11 [0.88;1.39] | 0.379 | 1.22 [0.95;1.58] | 0.132 | 1.32 [1.04;1.66] | 0.020 |
| No | Reference | | | | | | | | | | | |
| **Access to EHRs** | | | | | | | | | | | | |
| Yes | 1.67 [0.77;3.62] | 0.194 | 0.80 [0.39;1.66] | 0.554 | 0.97 [0.51;1.88] | 0.937 | .71 [0.41;1.22] | 0.215 | 0.72 [0.41;1.27] | 0.257 | 0.54 [0.29;0.99] | 0.045 |
| No | Reference | | | | | | | | | | | |
| **Duration of use of EHRs** | | | | | | | | | | | | |
| Only after COVID-19 outbreak | 0.07 [0.03;0.18] | <0.001 | 0.10 [0.04;0.25] | <0.001 | 0.56 [0.40;0.78] | 0.001 | 0.46 [0.34;0.62] | <0.00<br>1 | 0.57 [0.40;0.80] | 0.001 | 0.75 [0.56;1.01] | 0.060 |
| Before COVID-19 outbreak, but <2 years | 0.10 [0.06;0.17] | <0.001 | 0.12 [0.08;0.20] | 0.491 | 0.85 [0.60;1.19] | 0.336 | 0.53 [0.40;0.71] | <0.00<br>1 | 0.71 [0.51;0.97] | 0.032 | 0.817 [0.61;1.09] | 0.166 |
| [2-5[years | 0.23 [0.14;0.38] | <0.001 | 0.26 [0.18;0.38] | 0.807 | 0.87 [0.59;1.28] | 0.471 | 0.78 [0.56;1.07] | 0.121 | 1.172 [0.84;1.65] | 0.357 | 0.87 [0.63;1.21] | 0.414 |
| [5-10] years | 0.58 [0.34;0.99] | 0.047 | 0.51 [0.36;0.73] | | | | | | | | | |

| | | | | | | | | | | | | |
|---|---|---|---|---|---|---|---|---|---|---|---|---|
| > 10 years | Reference | | | **0.099** | 0.999<br>[0.99;1.01] | 0.580 | 1.00<br>[0.10;1.01] | 0.505 | 1.00<br>[0.10;1.01] | 0.307 | 1.00<br>[1.0;1.01] | 0.459 |
| **Frequency of access to EHRs** | | | | | | | | | | | | |
| Less than 1*month | 0.07<br>[0.03;0.12] | <0.001 | 0.15<br>[0.06;0.40] | 0.381 | 1.17<br>[0.83;1.65] | 0.383 | 0.80<br>[0.60;1.08] | 0.149 | 1.09<br>[0.78;1.52] | 0.620 | 0.93<br>[0.69;1.27] | 0.659 |
| At least 1* month | 0.12<br>[0.03;0.44] | 0.001 | 0.22<br>[0.06;0.75] | | | | | | | | | |
| At least 1* week | 0.16<br>[0.06;0.41] | <0.001 | 0.29<br>[0.12;0.69] | | | | | | | | | |
| More than 1* week | 0.20<br>[0.11;0.38] | <0.001 | 0.31<br>[0.18;0.54] | 0.142 | 0.77<br>[0.60;1.01] | **0.055** | 1.01<br>[0.81;1.26] | 0.964 | 0.83<br>[0.65;1.07] | 0.152 | 0.86<br>[0.69;1.08] | 0.862 |
| Everyday | Reference | | | | | | | | | | | |
